# The relationship between sleep disturbance and cognitive impairment in mood disorders: A systematic review

**DOI:** 10.1101/2021.08.12.21261470

**Authors:** Oliver Pearson, Nora Uglik-Marucha, Kamilla W. Miskowiak, Scott A. Cairney, Ivana Rosenzweig, Allan H. Young, Paul R.A. Stokes

**Affiliations:** Department of Biostatistics and Health Informatics, Institute of Psychiatry, Psychology and Neuroscience, King’s College London, SE5 8AF, London, UK; Centre for Affective Disorders, Department of Psychological Medicine, Institute of Psychiatry & Psychology and Neuroscience, King’s College London; Copenhagen Affective Disorder Research Centre (CADIC), Psychiatric Centre Copenhagen, Copenhagen University Hospital, Rigshospitalet, Copenhagen, Denmark; Department of Psychology, University of York, York, YO10 5DD; York Biomedical Research Institute, University of York, York, YO10 5DD; Sleep and Brain Plasticity Centre, Department of Neuroimaging, IoPPN, KCL; Sleep Disorders Centre, Guy’s and St Thomas’ Hospital, GSTT NHS, London; South London and Maudsley NHS Foundation Trust, Bethlem Royal Hospital, Monks Orchard Road, Beckenham, Kent, BR3 3BX, United Kingdom

**Author notes:** **Corresponding Author**: Oliver Pearson, Department of Biostatistics and Health Informatics, Institute of Psychiatry, Psychology & Neuroscience, King’s College London, 16 De Crespigny Park, London, SE5 8AF, UK.

**Keywords:** sleep disturbance, sleep disorder, insomnia, hypersomnia, cognitive impairment, bipolar disorder, major depressive disorder

## Abstract

**Background:** Cognitive impairment experienced by people with bipolar disorders (BD) or major depressive disorder (MDD) is associated with impaired psychosocial function and poorer quality of life. Sleep disturbance is another core symptom of mood disorders which may be associated with, and perhaps worsen, cognitive impairments. The aim of this systematic review was to critically assess the relationship between sleep disturbance and cognitive impairment in mood disorders.

**Methods:** In this systematic review, relevant studies were identified using electronic database searches of PsychINFO, MEDLINE, Embase and Web of Science.

**Findings:** Fourteen studies were included; eight investigated people with BD, five investigated people with MDD, and one included both people with MDD and people with BD. One study was an intervention for sleep disturbance and the remaining thirteen studies used either a longitudinal or cross-sectional observational design. Ten studies reported a significant association between subjectively measured sleep disturbance and cognitive impairment in people with MDD or BD after adjusting for demographic and clinical covariates, whereas no such association was found in healthy participants. Two studies reported a significant association between objectively measured sleep abnormalities and cognitive impairment in mood disorders. One study of cognitive behavioural therapy for insomnia modified for BD (CBTI-BD) found an association between improvements in sleep and cognitive performance in BD.

**Interpretation:** There is preliminary evidence to suggest a significant association between sleep disturbance and cognitive impairment in mood disorders. These findings suggest that identifying and treating sleep disturbance may be important when addressing cognitive impairment in mood disorders.

## Introduction

Cognitive impairment in people with bipolar disorders (BD) or major depressive disorder (MDD) is associated with reduced functional capacity (McIntyre et al., 2013; O’Donnell et al., 2017), poor illness prognosis (Martino et al., 2017; Zhang et al., 2020), and poor quality of life (Cambridge et al., 2018; Depp et al., 2012). Meta-analyses have reported significant deficits in executive function, memory and attention in people with MDD currently experiencing a depressive episode, with effect sizes ranging from *d* = -0.32 to -0.97 (Rock et al., 2014; Snyder, 2013). In BD, a meta-analysis has reported moderate effect size deficits in executive function in people experiencing depressive episodes (*d* = -0.55) and large effect sizes in people experiencing manic episodes (*d* = -0.72) (Kurtz & Gerraty, 2009). However, cognitive impairment is not confined to mood episodes, but persists during euthymia in MDD and BD. For example, Rock et al. (2014) found moderate effect size deficits in executive function and attention in people with remitted MDD, while Semkovska et al. (2019) identified deficits in working memory and long-term memory. In BD, medium to large effect size deficits in executive function, verbal memory, sustained attention and psychomotor speed have been identified by a meta-analysis (Bora et al., 2009). Nevertheless, there is considerable heterogeneity in cognitive impairments across people with mood disorders and approximately 35% of people with MDD (Pu et al., 2018) and 20-40% of people with BD (Burdick et al., 2014; Jensen et al., 2016) experience moderate to severe impairment in several cognitive domains.

The aetiology of cognitive impairment in mood disorders is not fully understood (Depp et al., 2016), and determining how cognitive impairment is mediated in MDD and BD is a key priority. Sleep disturbance may be one contributing factor. Sleep disturbances are core features of manic episodes, hypomanic episodes and major depressive episodes manifesting as insomnia or hypersomnia and fatigue in major depressive episodes and a decreased need for sleep in manic or hypomanic episodes (American Psychiatric Association, 2013). There is also growing evidence to suggest that sleep disturbance persists during euthymia in a substantial proportion of people with mood disorders. Clinically significant sleep disturbances are reported by 70% of euthymic people with BD (Harvey et al., 2005) and 44-60% of euthymic people with MDD (Carney et al., 2007; Iovieno et al., 2011; Nierenberg et al., 1999). Moreover, sleep disturbance is associated with higher rates of relapse and suicide attempts in both BD (Sylvia et al., 2012) and MDD (Li et al., 2012). Sleep disturbances escalate just before manic and depressive relapse in BD, and worsen during these episodes (Jackson et al., 2003).

There is strong evidence detailing the negative effects of sleep disturbance on many aspects of cognition in healthy individuals (Krause et al., 2017), including fragmented memory loss (Ashton et al., 2020) and impaired suppression of unwanted thoughts (Harrington et al., 2021; Harrington and Cairney, 2021). Recently, evidence has emerged to suggest an association between sleep disturbance and cognitive impairment in mood disorders. For example, Bradley et al. (2020) found that people with BD and abnormal sleep patterns, as measured by sleep actigraphy, performed significantly poorer than healthy participants in cognitive performance, whereas people with BD and normal sleep patterns did not significantly differ from healthy participants. As yet, there has not been a systematic review of the relationship between sleep disturbance and cognitive impairment in mood disorders.

The aim of this systematic review was to critically analyse the relationship between sleep disturbance and cognitive impairment in MDD and BD, and to examine relevant factors affecting this relationship.

## Methods

### Literature search

Two reviewers (OP and NUM) independently performed the literature search. Relevant studies were identified using electronic database searches of PsychINFO, MEDLINE, Embase and Web of Science. The following search string was used: [(sleep disturb*) OR (sleep disorder*) OR (insomn*) OR (hypersomn*)] AND [(cognitive impair*)] AND [(bipolar*) OR (depress*)]. Papers were extracted from inception until February 2021. Only studies written in English were considered. The relevance of the article was initially verified by title and abstract review and then by a further review of each manuscript to verify whether they met the inclusion and exclusion criteria. This systematic review followed PRISMA 2020 guidelines (Page et al., 2021), although a study protocol was not registered.

### Inclusion and exclusion criteria

All databases were screened following the same protocol. Included studies were required to include participants with a lifetime diagnosis of MDD, BD-I or BD-II, meeting criteria for ICD-9 (World Health Organization, 1988) or subsequent editions, or DSM-III (American Psychiatric Association, 1980) or subsequent editions. Included studies were required to measure both sleep disturbance and cognition. Studies were excluded if they: did not primarily include people with a diagnosis of MDD, BD-I or BD-II; included only participants with dysthymia or cyclothymia or with comorbid neurological disorders (e.g. dementia, epilepsy or stroke) or mild cognitive impairment (MCI); included primarily older participants; were follow-up studies or case reports; were not published by a peer-reviewed scientific journal.

### Data extraction

The following data were extracted: study name and year; participant sample size, age range, disorder and mood state of participants, measures of sleep and cognition, a summary of main findings and study limitations.

### Quality assessment

Two reviewers (OP and NUM) independently assessed the quality of included studies using the Quality Assessment Tool for Quantitative Studies, developed by the Effective Public Health Practice Project (EPHPP) (Thomas et al., 2003). The following criteria of each study were rated as either strong, moderate or weak: selection bias, design, confounders, blinding, data collection methods, and withdrawals/drop-outs (if applicable). These ratings were aggregated to form a global rating: studies were rated globally as strong if they received no weak ratings; moderate if they received one weak rating; and weak if they received two or more weak ratings.

## Results

A total of 3094 studies were identified, which was reduced to 2208 after duplicates were removed. Following primary assessment of titles and abstracts, this was reduced to 30 studies. Based on secondary assessment of full texts, we included 14 studies that fulfilled the eligibility criteria (please see Figure 1 for further details).

**Figure 1.**
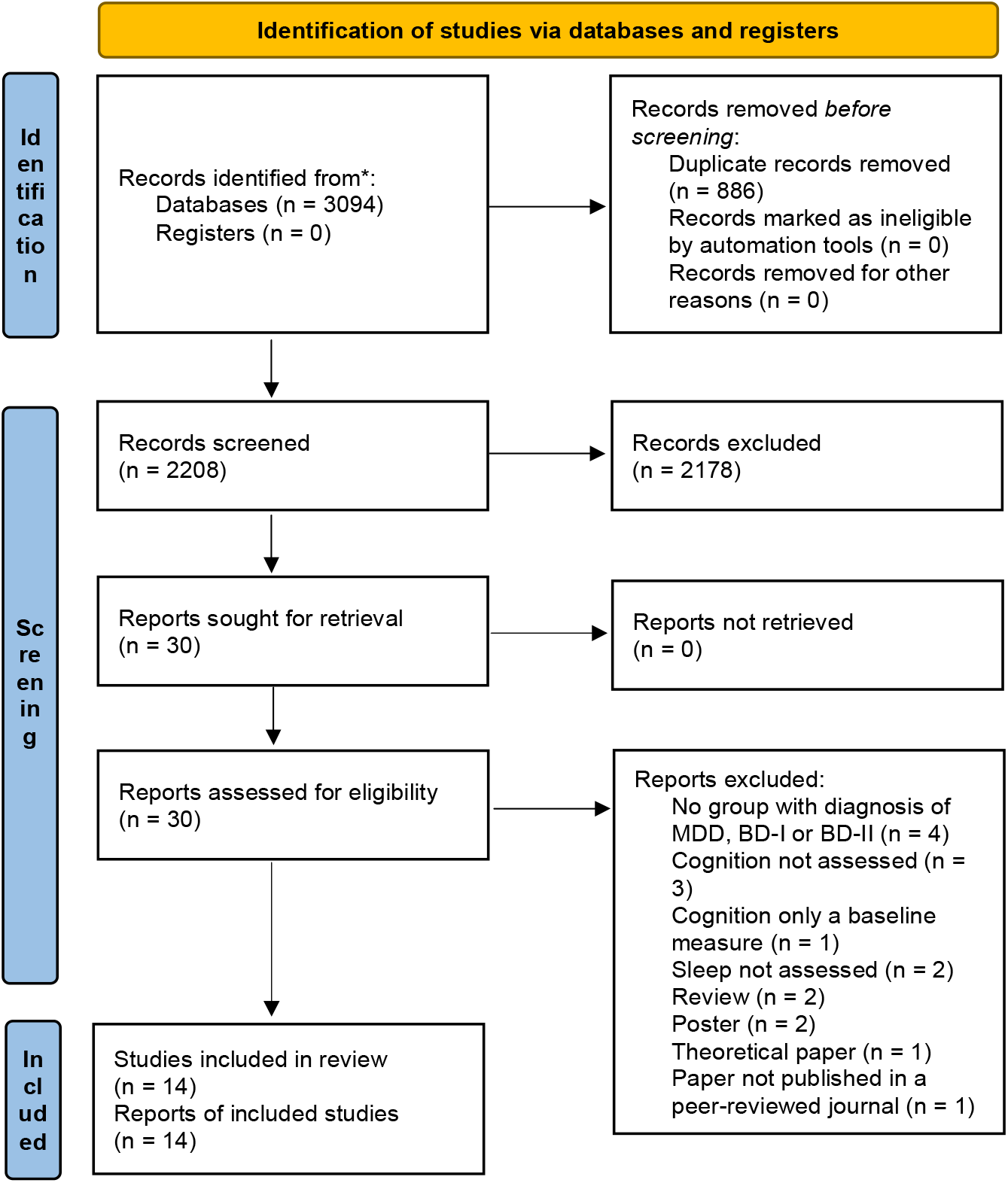
PRISMA 2020 Flow Diagram of study selection process

### Studies

Of the 14 included studies, eight included people with BD (Boland et al., 2015; Bradley et al., 2020; Kanady et al., 2017; Laskemoen et al., 2019; Samalin et al., 2017; Ullah, 2017; Volkert et al., 2015), five included people with MDD (Cabanel et al., 2019; Cha et al., 2019; Müller et al., 2017; Peng et al., 2018; Wilckens et al., 2020), and one included both people with MDD and people with BD (Soehner & Harvey, 2012) (see *Table 1*). These studies were published between years 2012-2020. A total of 2353 people with mood disorders were included in these studies, 1083 people with MDD and 1270 people with BD. The mood state of participants varied across studies. Of the eight BD studies, six included only euthymic participants (Boland et al., 2015; Kanady et al., 2017; Russo et al., 2015; Samalin et al., 2017, 2017; Volkert et al., 2015), whereas two included participants in any mood state (Bradley et al., 2020; Laskemoen et al., 2019). Of the five MDD studies, four only included people currently experiencing a major depressive episode (MDE); however, Wilckens et al. included only people with remitted depression. The one study of people with MDD or BD included participants in any mood state (Soehner & Harvey, 2012).

**Table 1.**
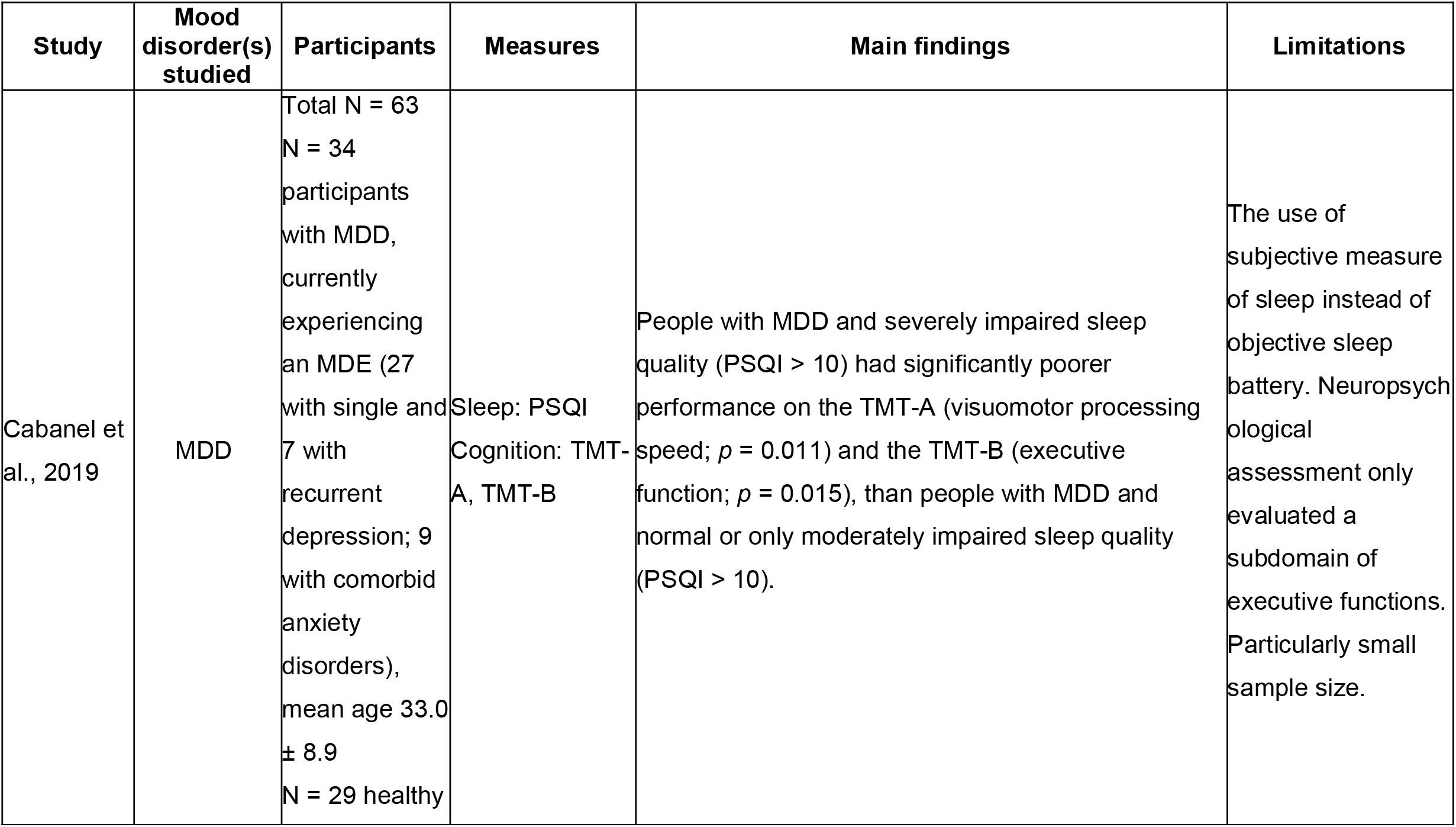
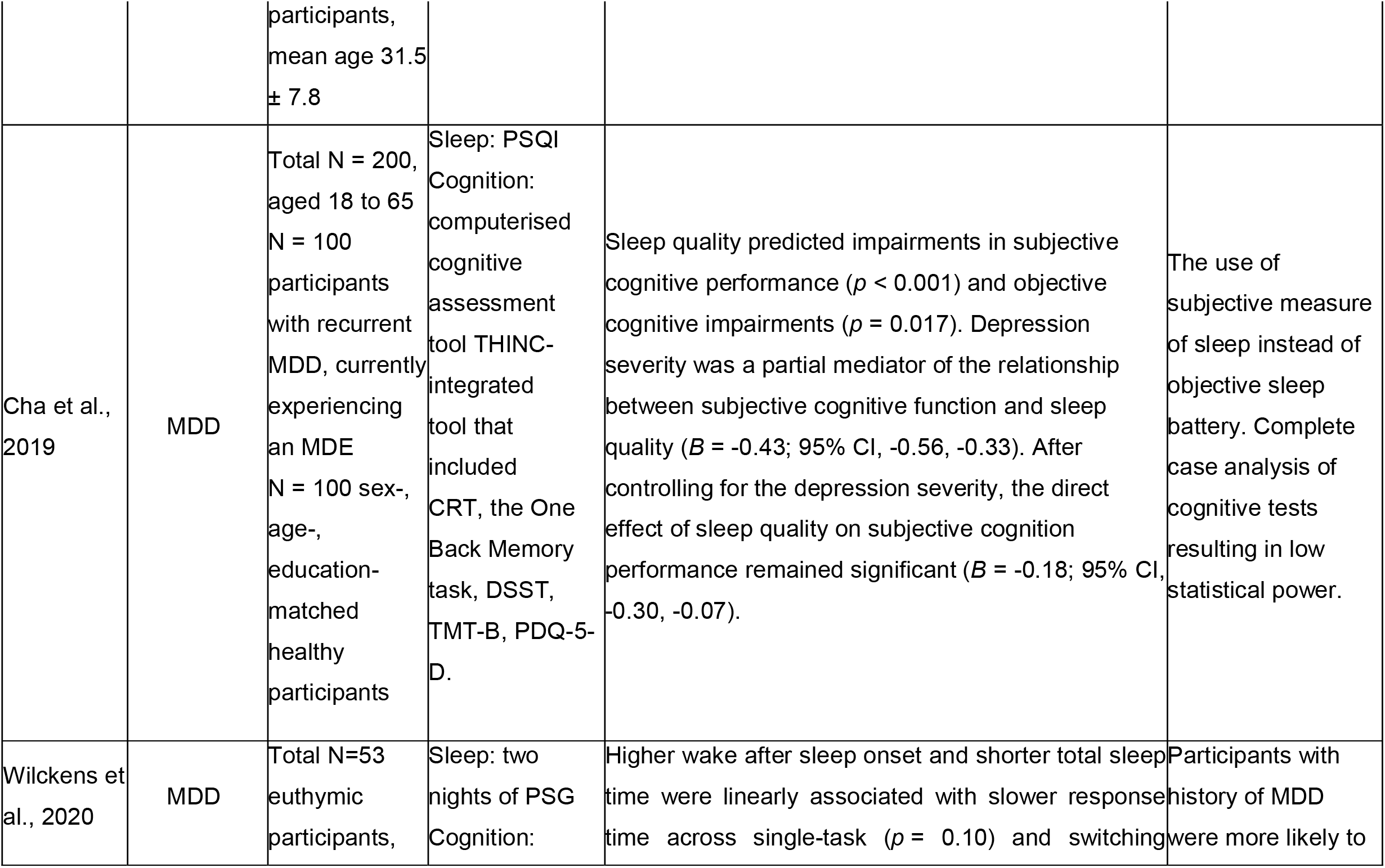
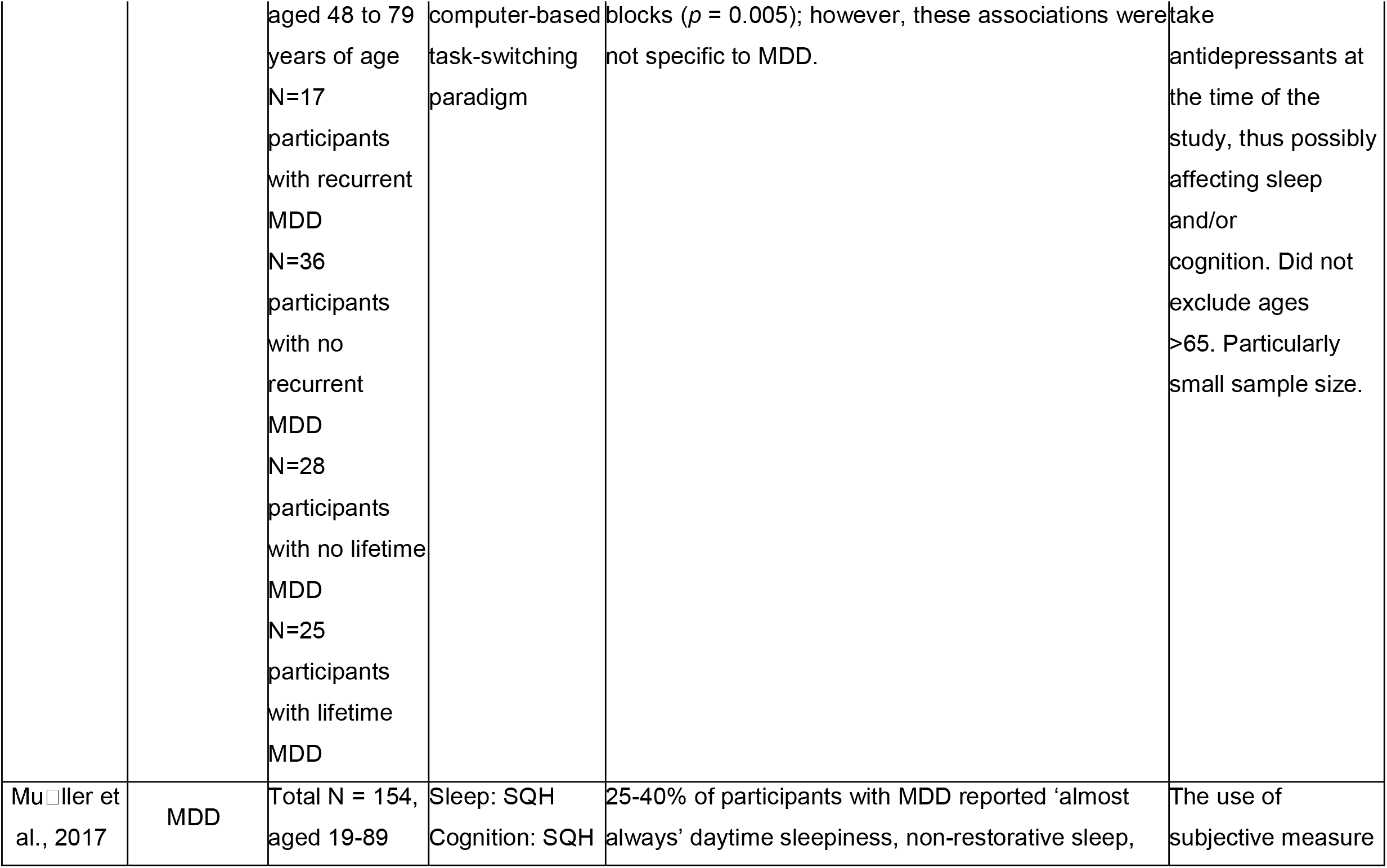
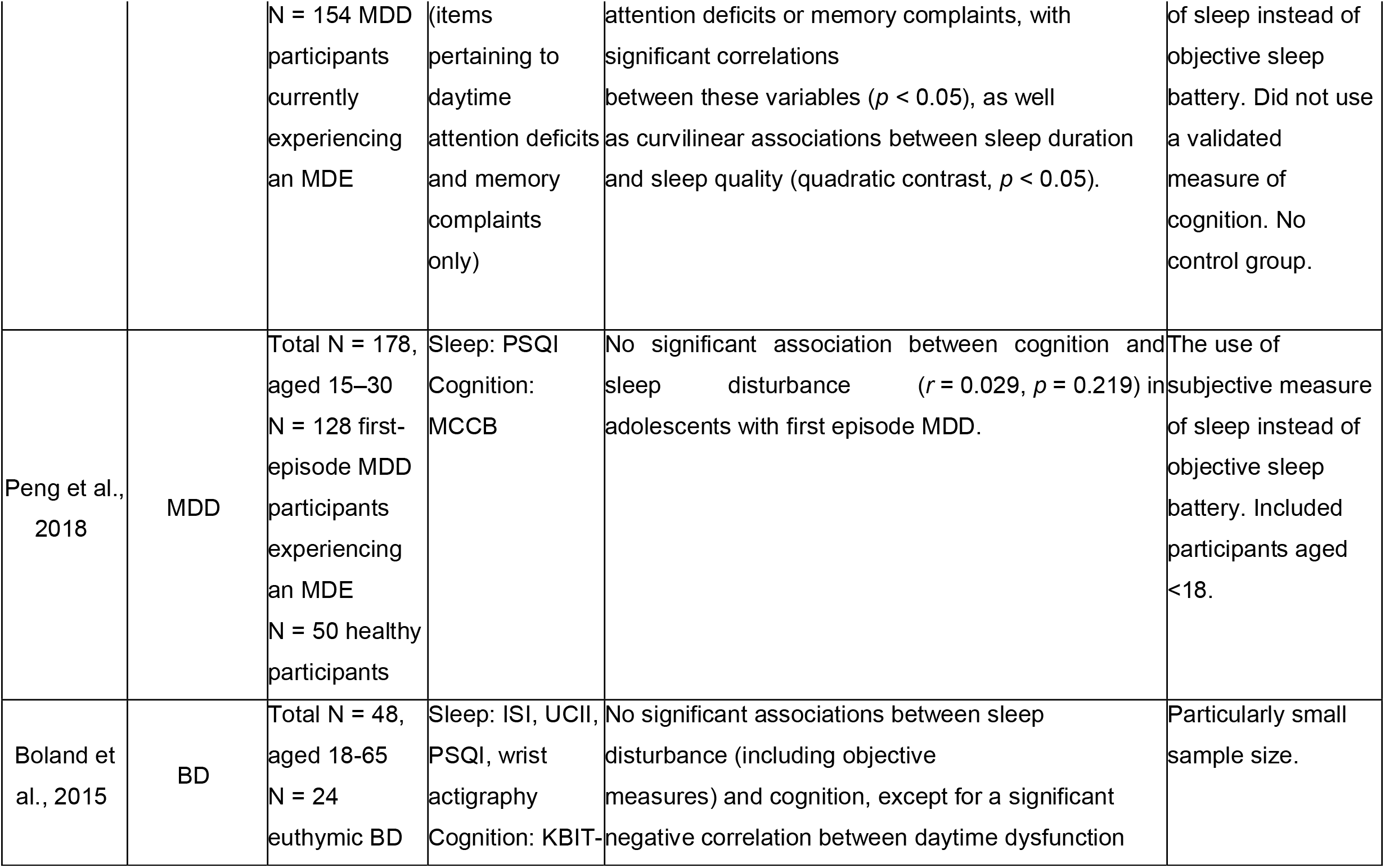
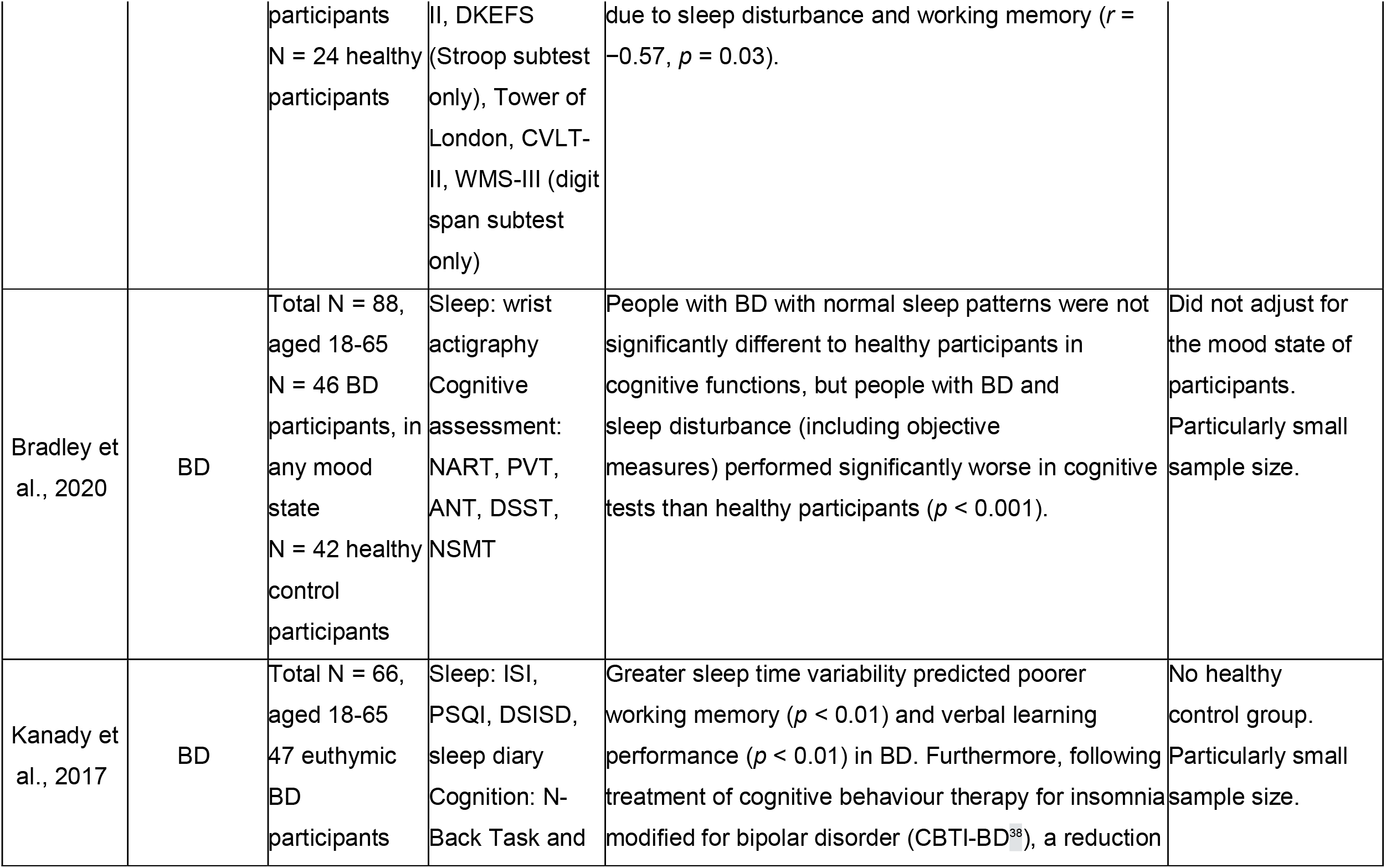
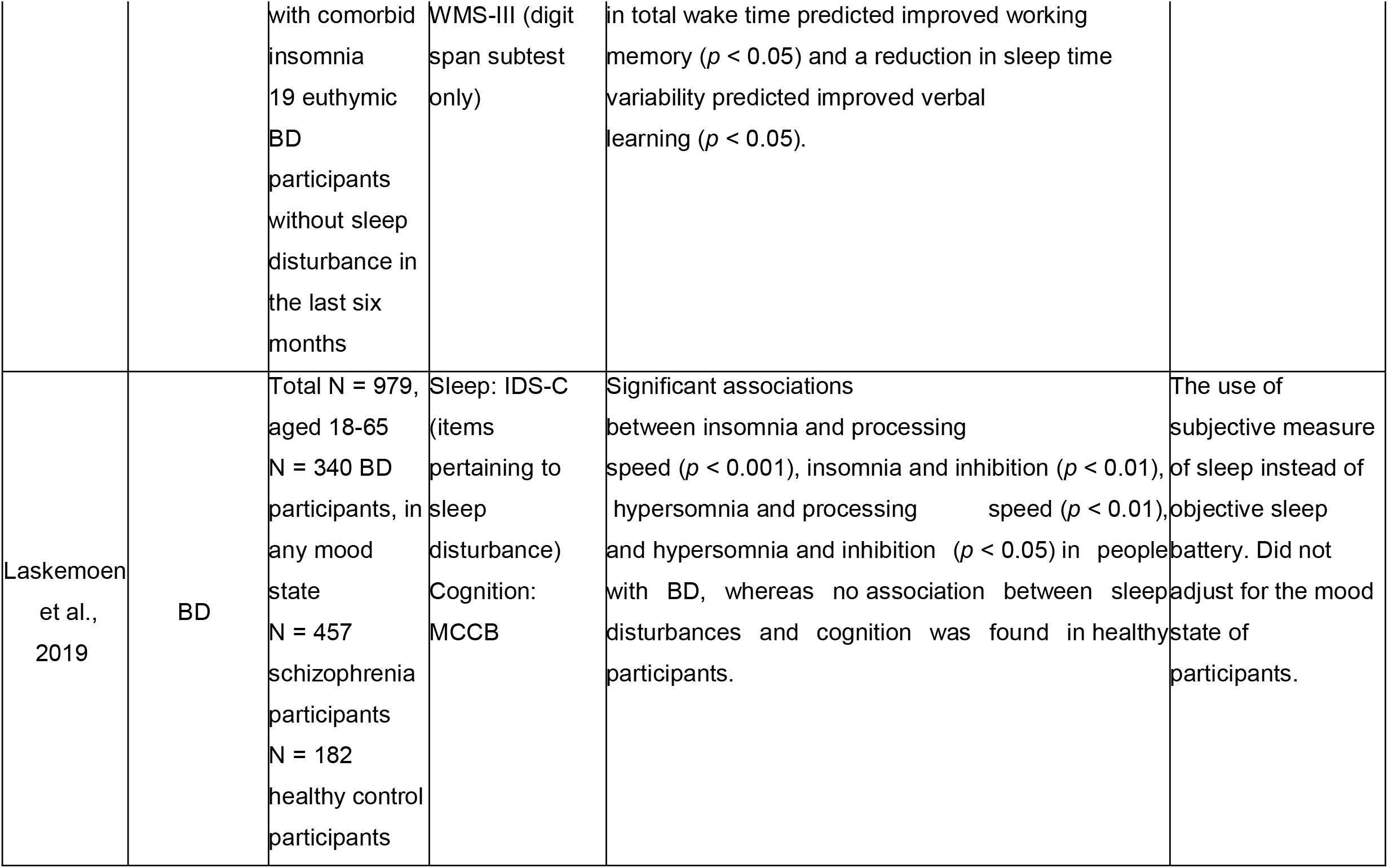
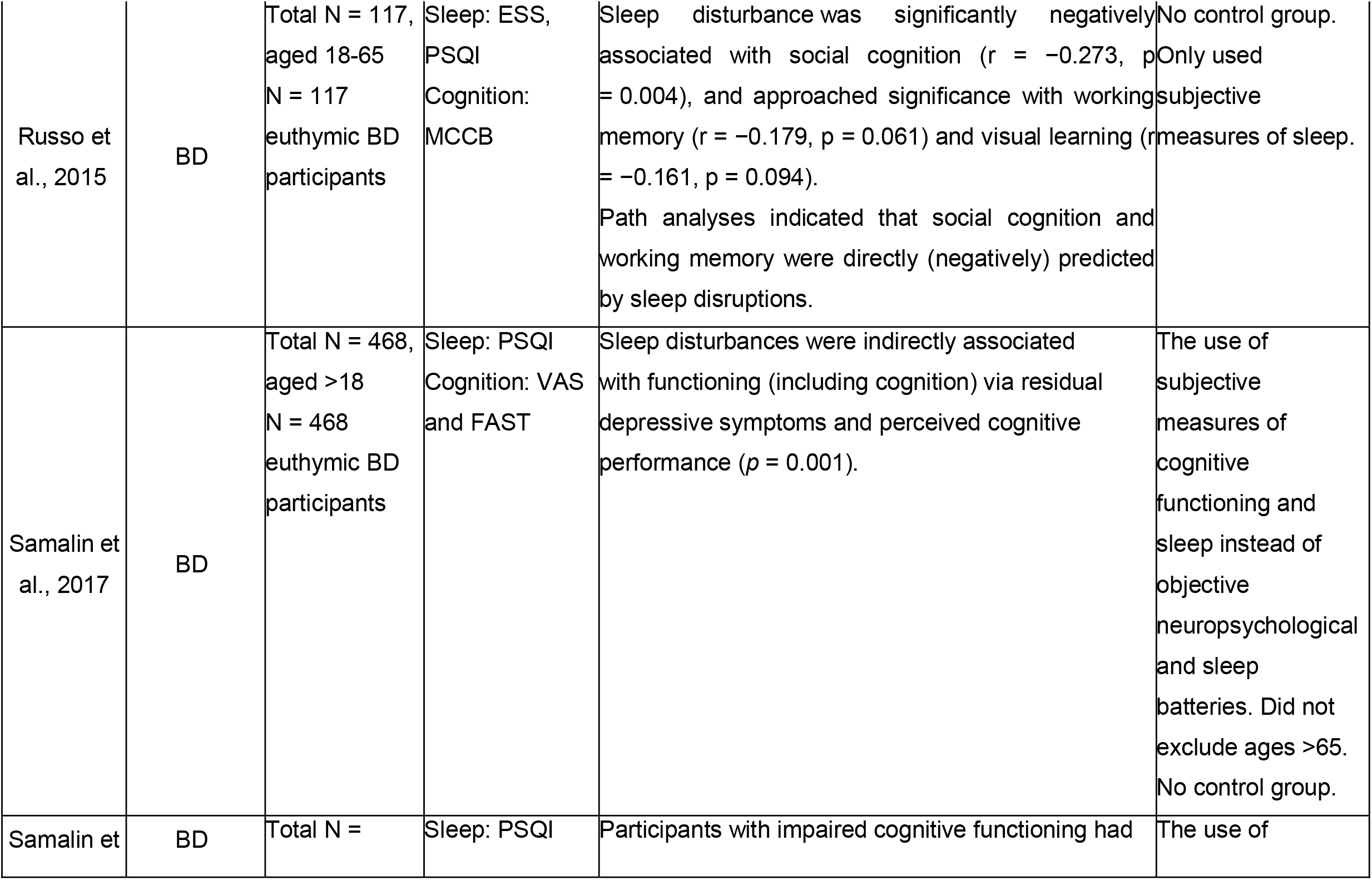
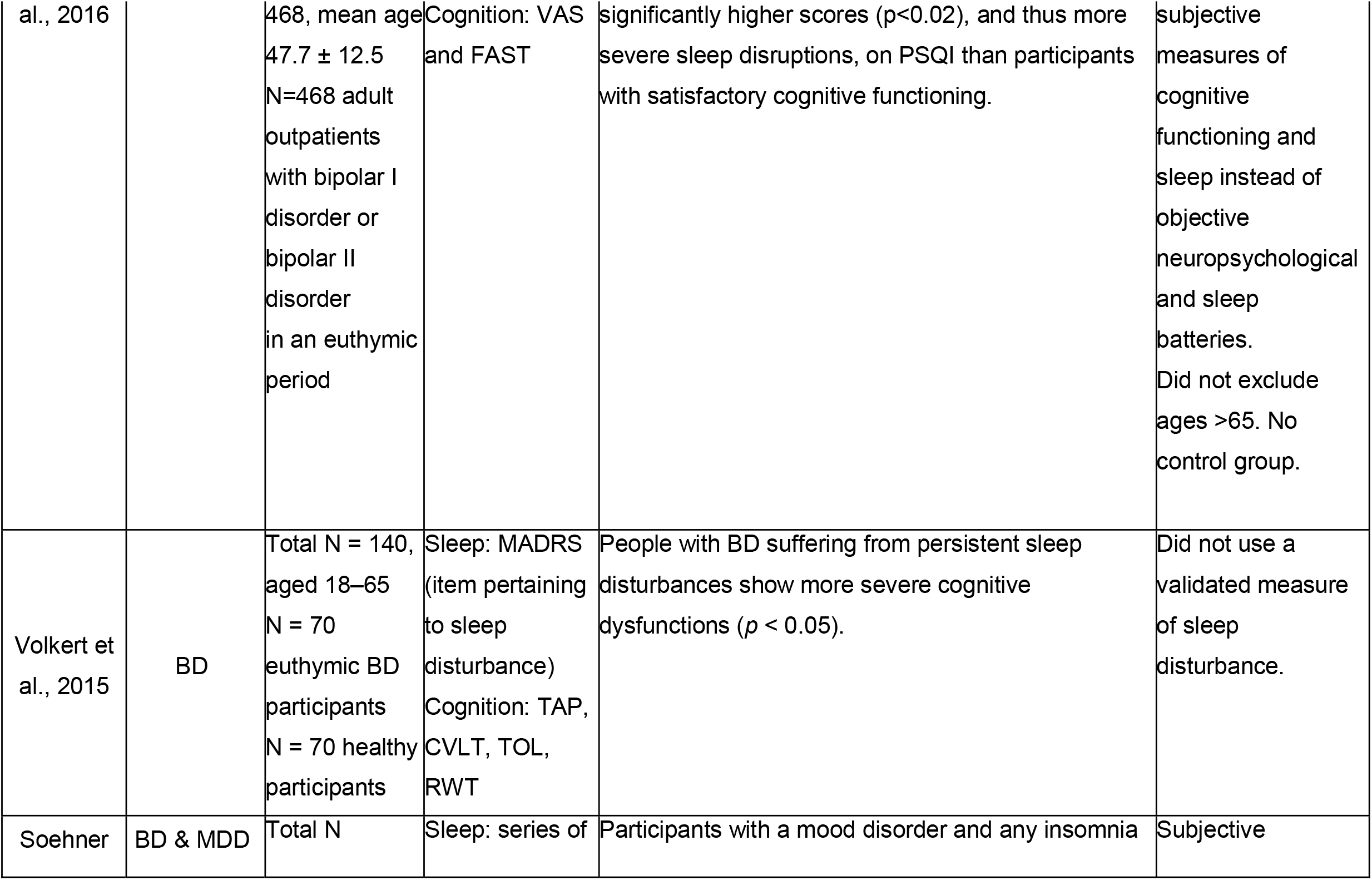
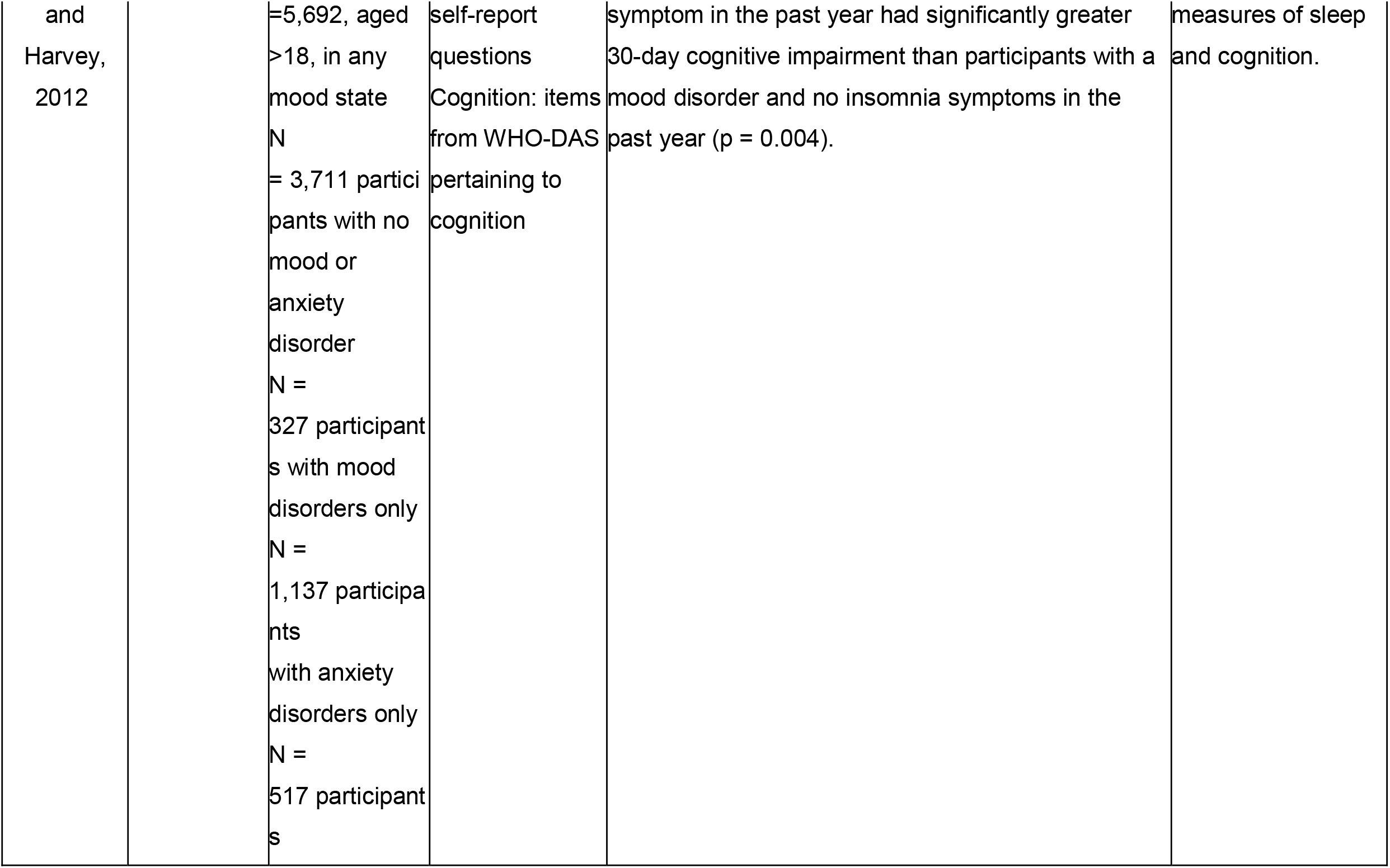
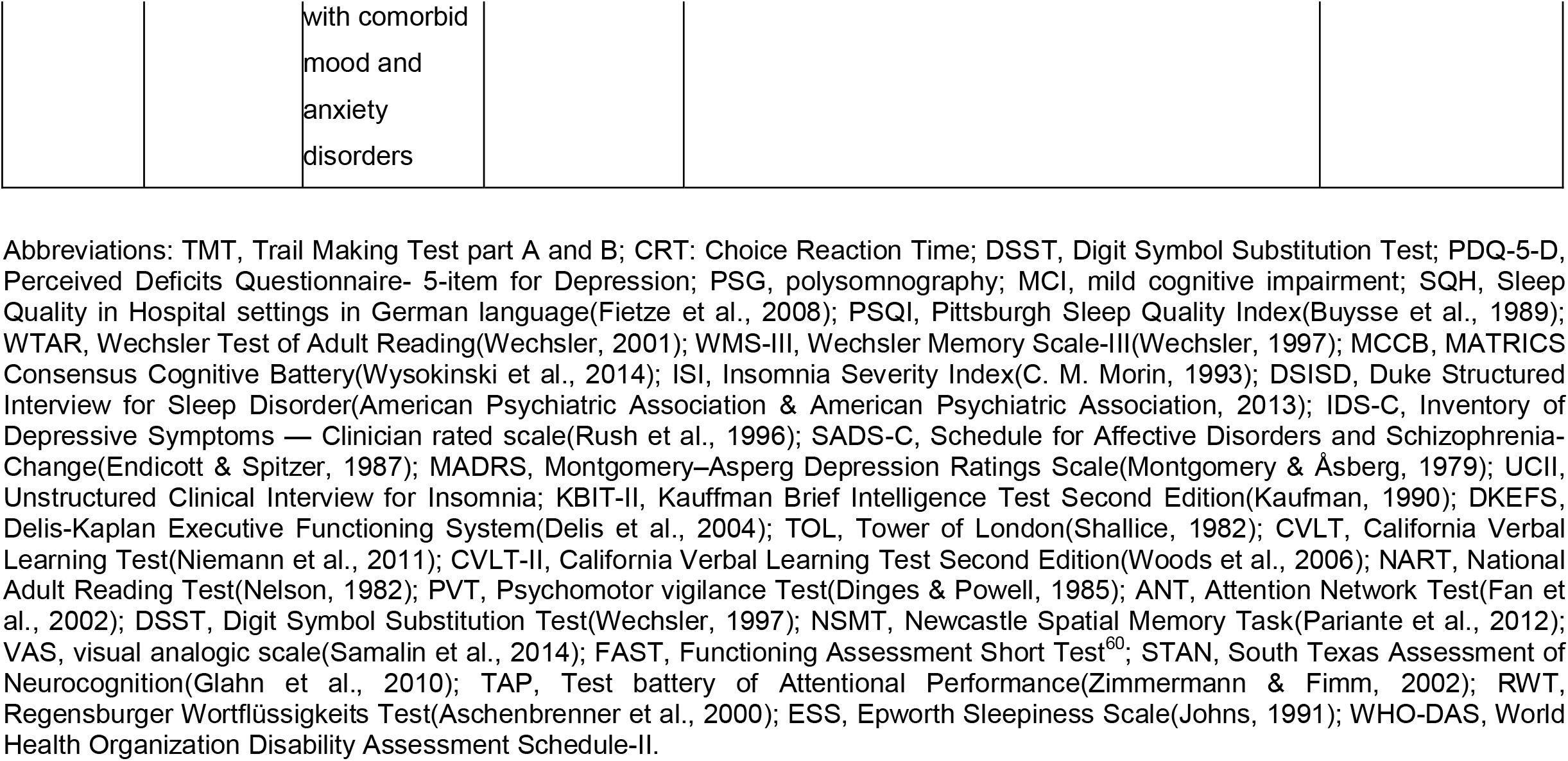
Characteristics and findings of studies examining sleep disturbance and cognitive impairment in MDD

### BD results

Six of the eight BD studies reported associations between sleep disturbance and cognitive impairment. Kanady et al. (2017) found that greater sleep time variability predicted poorer working memory (*p* < 0.01) and verbal learning performance (*p* < 0.01) in euthymic BD participants. Furthermore, following treatment with cognitive behaviour therapy for insomnia modified for bipolar disorder (CBTI-BD; Harvey et al., 2015), a reduction in total wake time predicted improved working memory (*p* < 0.05) and a reduction in sleep time variability predicted improved verbal learning (*p* < 0.05). Laskemoen et al. (2019) found significant associations between insomnia and processing speed (*p* < 0.001), insomnia and inhibition (*p* < 0.01), hypersomnia and processing speed (*p* < 0.01), and hypersomnia and inhibition (*p* < 0.05) in people with BD, in any mood state, whereas no association between sleep disturbances and cognition was found in healthy participants. Samalin et al. (2017) found that sleep disturbances were indirectly associated with cognitive function potentially mediated via residual depressive symptoms and perceived cognitive performance, in euthymic BD participants (*p* = 0.001). Another study by Samalin et al. (2016) reported that euthymic BD participants with impaired cognitive functioning had significantly higher PSQI scores (p<0.02), and thus more severe sleep disruptions, than participants with satisfactory cognitive functioning. Volkert et al. (2015) reported that euthymic BD participants experiencing persistent sleep disturbances show more severe cognitive dysfunctions (*p* < 0.05). Bradley et al. (2020) found that people with BD, in any mood state, with normal sleep patterns were not significantly different to healthy participants in cognitive functions, but people with BD and sleep disturbance (including objective measures) performed significantly worse in cognitive tests than healthy participants (*p* < 0.001). Russo et al. (2015) found that sleep disturbance was significantly negatively associated with social cognition (r = -0.273, p = 0.004), but only associated at a trend level with working memory (r = -0.179, p = 0.061) and visual learning (r = -0.161, p = 0.094), in euthymic BD participants. Boland et al. (2015) reported no significant associations between sleep disturbance (including objective parameters: wake after sleep onset, total sleep duration, and sleep efficiency) and cognition, except for a significant negative correlation between daytime dysfunction due to sleep disturbance and working memory (*r* = -0.57, *p* = 0.03), in euthymic BD participants.

### MDD results

Cabanel et al. (2019) reported that MDD participants experiencing an MDE with severely impaired sleep quality (PSQI > 10) had significantly poorer performance on the TMT-A (visuomotor processing speed; *p* = 0.011) and the TMT-B (executive function; *p* = 0.015) than those experiencing an MDE but with normal or moderately impaired sleep quality (PSQI < 10). In a study of MDD participants experiencing an MDE, Cha et al. (2019) found that sleep quality predicted impairments in subjective cognitive performance (*p* < 0.001) and objective cognitive impairments (*p* = 0.017). Depression severity was a partial mediator of the relationship between subjective cognitive function and sleep quality (*B* = -0.43; 95% CI, -0.56, -0.33; *B* = -0.18; 95% CI, -0.30, -0.07 after controlling for depression severity). In a study of euthymic people with remitted MDD, Wilckens et al. (2020) reported that higher wake after sleep onset (i.e., longer periods of wakefulness occurring after defined sleep onset), and shorter total sleep time, were linearly associated with slower response time across single-task (*p* = 0.10) and switching blocks (*p* = 0.005) considered a proxy for condition-independent psychomotor slowing (White et al., 1997); however, these associations were also observed in healthy controls and therefore not specific to MDD. Müller et al. (2017) found that 25-40% of MDD participants experiencing an MDE reported ‘almost always’ daytime sleepiness, non-restorative sleep, attention deficits or memory complaints, with significant correlations between these variables (*p* < 0.05), as well as curvilinear associations between sleep duration and sleep quality (quadratic contrast, *p* < 0.05). However, Peng et al. (2018) reported no significant association between cognition and sleep disturbance (*r* = 0.029, *p* = 0.219) in adolescents with first episode MDD, experiencing an MDE.

### Results in mixed MDD & BD groups

Soehner and Harvey (2012) (N = 639 MDD participants; N = 138 BD participants, in any mood state) reported that participants with a mood disorder and insomnia in the past year experienced significantly greater cognitive impairment in the past 30 days than participants with a mood disorder and no insomnia symptoms in the past year (p = 0.004).

### Methodological quality assessment

Methodological quality assessment of studies using the EPHPP Quality Assessment Tool for Quantitative Studies (Thomas et al., 2003) are displayed in *Supplementary Table 1*. Selection bias was generally rated as moderate because in most studies participants were sampled from either outpatient clinics or inpatient settings. Study design was generally rated as weak because most were cross-sectional. Most studies controlled for at least 80% of relevant confounding variables, such as age, coffee consumption etc. Blinding was rated as moderate if it was not described, as was the case in almost all studies. Most studies used validated and reliable measures for sleep disturbance (e.g. Pittsburgh Sleep Quality Index, PSQI; Buysse et al., 1989) and cognition (e.g. MATRICS Consensus Cognitive Battery, MCCB; Wysokinski et al., 2014), and were therefore rated strong. Where applicable, withdrawals were reported satisfactorily by most studies. There was no association between global rating and positivity of findings.

## Discussion

The purpose of this systematic review was to critically analyse the published literature investigating the relationship between sleep disturbance and cognitive impairment in BD and MDD. Fourteen studies were included in this review, for which quality assessment ranged from weak to strong.

Kanady et al. (2017) was the only RCT included in this review. This study, in which CBTI-BD (Harvey et al., 2015) was administered to participants with BD, found that a reduction in total wake time predicted improved working memory. The study also found that a reduction in sleep time variability predicted improved verbal learning. These findings suggest that sleep disturbance may contribute to cognitive impairment in BD, and that CBTI-BD may be an effective treatment. The remaining studies included in our review used either a longitudinal or cross-sectional design, and therefore did not allow for causal inferences to be made.

Ten of the twelve studies that measured sleep disturbance using subjective measures in participants with either MDD or BD found a significant association between sleep disturbance and cognitive impairment when adjusting for demographic and clinical covariates. In contrast, no significant association was reported in healthy participants, which suggests that this relationship may be specific to mood disorders. Given the strong evidence on the effect of objective sleep disturbance on cognitive impairment in healthy individuals (Krause et al., 2017), this may indicate that subjective experience of sleep impairment may also increase risks for cognitive impairment in people with mood disorders. Some studies also found a curvilinear association, indicating that both extremely low (<5 hours) and extremely high (>11 hours) self-reported sleep durations were associated with cognitive impairment; this was observed in participants with MDD (e.g. Müller et al., 2017) and in BD (e.g. Laskemoen et al., 2019). However, there was also evidence to suggest that insomnia may have more deleterious effects on cognitive impairment than hypersomnia. For example, Laskemoen et al. (2019) found a larger effect size between insomnia and processing speed (*F* = 15.43, *p* < 0.001, η^2^ = 0.019) than hypersomnia (*F* = 6.87, *p <* 0.01, η^2^ = 0.009) in people with BD.

The main disadvantage of using subjective measures to assess sleep disturbance in people with mood disorders is that there is a considerable risk of bias. Euthymic people with either BD or MDD tend to significantly underestimate sleep duration and quality (Harvey et al., 2005; Carney et al., 2011). Indeed, Boland et al. (2015) found that people with BD had significantly higher insomnia severity index (ISI) scores than healthy participants (indicating worse sleep quality), yet found no significant difference between BD and healthy participants in objective sleep parameters, as measured by actigraphy. This effect may be due to maladaptive sleep-related beliefs developed during mood episodes, in which sleep disturbance is particularly severe, which are then carried over into euthymia (Carney et al., 2011). Nevertheless, subjective measures provide valuable information about the perception of sleep disturbance and sleep-related cognitions, and interventions of maladaptive sleep beliefs may be a useful way of treating sleep disturbance in mood disorders (Morin et al., 2002). Although antidepressant treatment produces no change in maladaptive sleep beliefs in MDD (Carney et al., 2011), Cognitive Behavioural Treatment for Insomnia (CBT-I) has been shown to significantly improve maladaptive sleep beliefs in people with insomnia. CBT-I has been found to significantly improve both subjective sleep measures, measured by sleep diaries, and objective sleep measures, measured by polysomnography (PSG) (Morin et al., 2002). Therefore, sleep-related cognitions may be an important target for treating sleep disturbance and potentially cognitive impairment in people with mood disorders.

Only three studies included in this review measured sleep disturbance using objective measures: two examined sleep disturbance in participants with BD using actigraphy (Boland et al., 2015; Bradley et al., 2020) and one investigated sleep disturbance in participants with MDD using PSG (Wilckens et al., 2020). The MDD study that used PSG found that longer periods of wakefulness occurring after defined sleep onset were associated with psychomotor slowing, however these associations were not specific to MDD (Wilckens et al., 2020). One advantage of PSG is that it is more accurate than actigraphy in recognising sleep and wake times, and is therefore the ‘gold standard’ of objective sleep measurement. Another key advantage of PSG is the ability to measure sleep architecture, however, Wilckens et al. (2020) did not report sleep architecture measurements.

Mood disorders are associated with several sleep architecture abnormalities, particularly altered distribution of rapid eye movement (REM) sleep in MDD (Hutka et al., 2021) and BD (Eidelman et al., 2010). Components of sleep architecture have been linked to aspects of cognitive impairment in healthy individuals. For example, time in slow wave sleep contributes to processing speed in healthy individuals (della Monica et al., 2018). Abnormal sleep architecture may be associated with cognitive impairment in mood disorders, so sleep architecture may be a highly relevant variable to measure. Moreover, knowledge of the specific components of sleep archiecture associated with improvements in mental health or cognition could be targeted in interventions for treatment or prevention. For example, enhancing slow oscillation activity via non-invasive auditory stimulation has been shown to improve memory in healthy individuals (Ngo et al., 2013). Thus, we suggest that future studies of the role of sleep in cognition in mood disorders use PSG where possible. It is worth noting that there is a well-known ‘first night effect’ in PSG, characterised by decreased total sleep time, decreased sleep efficiency and reduction of REM sleep on the first night of measurement (Agnew, Webb, & Williams, 1966), so future PSG studies should ideally be conducted over multiple nights to allow for a more accurate assessment.

Interestingly, the two BD studies which used actigraphy had opposite findings. Bradley et al. (2020) found a significant association between objective sleep disturbance and cognitive impairment in participants with BD (*p* < 0.001). Specifically, participants with BD and abnormal sleep patterns were found to experience deficits in attention and executive function but normal verbal memory. In contrast, Boland et al. (2015) found no significant association between objective sleep disturbance and cognitive impairment in participants with BD. However, this study included a relatively small sample size of participants with BD (n=24) which implies low statistical power and risk of type-II error. Nevertheless, taken together, these two BD studies are inconclusive as to the association between objective sleep disturbance and cognitive impairment in people with BD and highlights the need for future research using larger sample sizes and PSG.

In studies examining sleep disturbance and cognitive impairment in mood disorders, it is normally best practice to include only euthymic participants because mood state may affect both sleep disturbance and cognitive impairment, and therefore may be a confound (Miskowiak et al., 2018). The MDD studies were particularly weak in this regard, as four of the five MDD studies only included people currently experiencing a depressive episode. The BD studies were stronger, but still varied: six studies only included participants that were euthymic (Boland et al., 2015; Kanady et al., 2017; Russo et al., 2015; Samalin et al., 2016, 2017; Volkert et al., 2015), and two included participants in any mood state (Bradley et al., 2020; Laskemoen et al., 2019). The study in mixed MDD & BD groups included participants in any mood state (Soehner and Harvey, 2012). In comparing across studies, it is apparent that mood state did not seem to significantly affect the relationship between sleep disturbance and cognitive impairment. In BD, of the six studies that reported significant associations, five included euthymic participants and one included participants in any mood state; and of the two that reported non-significant associations, one included euthymic participants and one included participants in any mood state. In MDD, of the four studies that reported significant associations, three included participants currently experiencing an MDE and one included euthymic participants with remitted MDD; and the one study that reported non-significant associations included participants currently experiencing an MDE. Therefore, there did not seem to be a clear trend between the mood state of participants and significance of results.

With the exception of Wilckens et al. (2020), none of the included studies adjusted for the use of psychotropic medications in the relationship between sleep disturbance and cognitive impairment. Psychotropic medication may be a confounding variable, as they can affect both sleep and cognition in people with mood disorders (Staner, 2005). Indeed, Volkert et al. (2015) reported that antipsychotic treatment and polypharmacy were related to cognitive impairment. Furthermore, many antipsychotics affect REM sleep, which has been linked to emotional memory processing (Walker & van Der Helm, 2009). Without adjusting for the use of psychotropic medications, it is unclear which medications alter the relationship between sleep disturbance and cognitive impairment in mood disorders, and to what extent.

### Limitations

This review reflects the limited number of studies in the field and so the relatively small number of participants in which the associations between sleep disturbance and cognitive impairment have been investigated, particularly regarding MDD, limits our power to draw wider conclusions. The studies included in our review generally had small sample sizes (*N* = 48 - 4555), which limited statistical power and increased risks of type-II error. A meta-analysis was not performed because there was significant heterogeneity between studies in the age and mood state of participants, as well as the measures of sleep and cognition used.

### Suggestions for future research

Theoretically, sleep disturbance may cause, predispose or exacerbate cognitive impairment in mood disorders but this link has yet to be conclusively evaluated. Further research is clearly needed in this area in participants with mood disorders. Based on the evaluation of current evidence, we would suggest that future studies follow five recommendations. First, studies should include only euthymic participants, to remove the confounding effect of mood state. Second, they should use objective measures of sleep, ideally PSG, to evaluate the role of sleep disturbance in mediating cognitive impairment, and to determine which aspects of sleep architecture are linked to cognition in these individuals. Third, all studies should use a comprehensive cognitive battery such as the BACS-A (Keefe et al., 2004) or ideally the ISBD-BANC (Yatham et al., 2010). Fourth, analyses should adjust for the use of psychotropic medications, as these may also have a confounding effect. Finally, we would suggest that studies use a prospective longitudinal design, and, if possible, use a randomised clinical trial design for intervention studies.

### Conclusion

In conclusion, our systematic review provides preliminary evidence for an association between sleep disturbance and cognitive impairment in mood disorders. The studies identified by this systematic review found increased rates of both subjective sleep disturbances and cognitive impairment in people with MDD and BD, compared to healthy participants. Ten of the twelve studies that measured sleep disturbance using *subjective* measures of sleep found a significant association between sleep disturbance and cognitive impairment across both diagnoses when adjusting for demographic and clinical covariates, whereas no significant association was found in healthy participants. In contrast, the evidence was scarcer and more conflicting with regards to whether *objectively* measured sleep abnormalities are associated with cognitive impairment in people with mood disorders: two out of three studies reported significant associations. Only one study (Kanady et al., 2017) conducted an intervention, and reported that improved sleep predicted an improvement in cognition in BD. As cognitive impairment has significant implications for psychosocial functioning in people with mood disorders, these findings highlight the importance of effectively identifying and treating sleep disturbances in people with MDD and BD.

## Data Availability

No data was used.

## Contributors

OP wrote the report with input from NUM, KWM, SAC, IR, AHY and PRAS. OP and NUM independently performed the literature search and quality assessment. All authors had full access to all the papers in the study and had final responsibility for the decision to submit for publication.

Conceptualisation: OP and PRAS, data curation: n/a, formal analysis: n/a, funding acquisition: n/a, investigation: OP and NUM, methodology: n/a, project administration: OP and PRAS, resources: n/a, software: n/a, supervision: PRAS, validation: OP, visualisation: OP, writing – original draft: OP, writing – review & editing: OP, NUM, KWM, SAC, IR, AHY and PRAS.

## Declaration of interests

OP, NUM, SAC and IR declare no competing interests.

KWM has received consultancy fees from Lundbeck and Janssen-Cilag in the past three years.

PRAS reports non-financial support from Janssen Research and Development LLC, personal fees and non-financial support from Frontiers in Psychiatry, personal fees from Allergan, a grant from H Lundbeck, grants and non-financial support from Corcept Therapeutics, outside the submitted work.

AHY declares:

Employed by King’s College London; Honorary Consultant SLaM (NHS UK)

Deputy Editor, BJPsych Open

Paid lectures and advisory boards for the following companies with drugs used in affective and related disorders:

Astrazenaca, Eli Lilly, Lundbeck, Sunovion, Servier, Livanova, Janssen, Allegan, Bionomics, Sumitomo Dainippon Pharma, COMPASS

Consultant to Johnson & Johnson

Consultant to Livanova

Received honoraria for attending advisory boards and presenting talks at meetings organised by LivaNova. Principal Investigator in the Restore-Life VNS registry study funded by LivaNova.

Principal Investigator on ESKETINTRD3004: “An Open-label, Long-term, Safety and Efficacy Study of Intranasal Esketamine in Treatment-resistant Depression.”

Principal Investigator on “The Effects of Psilocybin on Cognitive Function in Healthy Participants”

Principal Investigator on “The Safety and Efficacy of Psilocybin in Participants with Treatment-Resistant Depression (P-TRD)”

UK Chief Investigator for Novartis MDD study MIJ821A12201

Grant funding (past and present): NIMH (USA); CIHR (Canada); NARSAD (USA); Stanley Medical Research Institute (USA); MRC (UK); Wellcome Trust (UK); Royal College of Physicians (Edin); BMA (UK); UBC-VGH Foundation (Canada); WEDC (Canada); CCS Depression Research Fund (Canada); MSFHR (Canada); NIHR (UK). Janssen (UK)

No shareholdings in pharmaceutical companies

## Supplementary Tables

**Supplementary Table 1:** Methodological evaluation of studies using the EPHPP Quality Assessment Tool for Quantitative Studies

| **Author and year** | **Mood disorder(s) studied** | **Selection bias** | **Study design** | **Confounders** | **Blinding** | **Data collection method** | **Withdrawals and dropouts** | **Global rating** |
| --- | --- | --- | --- | --- | --- | --- | --- | --- |
| Soehner and Harvey, 2012 | MDD and BD | Moderate | Weak | Strong | Moderate | Moderate | N/A | **Moderate** |
| Cabanel et al., 2019 | MDD | Moderate | Weak | Strong | Moderate | Strong | N/A | **Moderate** |
| Cha et al., 2019 | MDD | Moderate | Weak | Strong | Moderate | Strong | Strong | **Moderate** |
| Muller et al., 2017 | MDD | Moderate | Weak | Strong | Moderate | Weak | N/A | **Weak** |
| Peng et al., 2018 | MDD | Moderate | Weak | Strong | Moderate | Strong | N/A | **Moderate** |
| Wilckens et al., 2020 | MDD | Moderate | Weak | Strong | Moderate | Strong | N/A | **Moderate** |
| Boland et al., 2015 | BD | Moderate | Moderate | Strong | Strong | Strong | Weak | **Moderate** |
| Bradley et al., 2020 | BD | Strong | Moderate | Strong | Moderate | Strong | Strong | **Strong** |
| Kanady et al., 2017 | BD | Moderate | Strong | Strong | Moderate | Strong | Strong | **Strong** |
| Laskemoen et al., 2020 | BD | Strong | Weak | Strong | Moderate | Moderate | N/A | **Moderate** |
| Russo et al., 2015 | BD | Moderate | Weak | Moderate | Moderate | Strong | N/A | **Moderate** |
| Samalin et al., 2017 | BD | Strong | Weak | Weak | Moderate | Strong | N/A | **Weak** |
| Samalin et al., 2016 | BD | Strong | Weak | Weak | Moderate | Strong | N/A | **Weak** |
| Volkert et al., 2015 | BD | Moderate | Weak | Strong | Moderate | Moderate | N/A | **Moderate** |

